# Basal ganglia theta power indexes trait anxiety in people with Parkinson’s disease

**DOI:** 10.1101/2024.06.04.24308449

**Authors:** Bart E.K.S. Swinnen, Colin W. Hoy, Elena Pegolo, Elena Ubeda Matzilevich, Julia Sun, Bryony Ishihara, Francesca Morgante, Erlick Pereira, Fahd Baig, Michael Hart, Huiling Tan, Zimi Sawacha, Martijn Beudel, Sarah Wang, Philip Starr, Simon Little, Lucia Ricciardi

## Abstract

**Background:** Neuropsychiatric symptoms are common and disabling in Parkinson’s disease (PD), with troublesome anxiety occurring in one-third of patients. Management of anxiety in PD is challenging, hampered by insufficient insight into underlying mechanisms, lack of objective anxiety measurements, and largely ineffective treatments.

In this study, we assessed the intracranial neurophysiological correlates of anxiety in PD patients treated with deep brain stimulation (DBS) in the laboratory and at home. We hypothesized that low-frequency (theta-alpha) activity would be associated with anxiety.

**Methods:** We recorded local field potentials (LFP) from the subthalamic nucleus (STN) or the globus pallidus pars interna (GPi) DBS implants in three PD cohorts: 1) patients with recordings (STN) performed in hospital at rest via perioperatively externalized leads, without active stimulation, both ON or OFF dopaminergic medication; 2) patients with recordings (STN or GPi) performed at home while resting, via a chronically implanted commercially available sensing-enabled neurostimulator (Medtronic Percept^TM^ device), ON dopaminergic medication, with stimulation both ON or OFF; 3) patients with recordings performed at home while engaging in a behavioral task via STN and GPi leads and electrocorticography paddles (ECoG) over premotor cortex connected to an investigational sensing-enabled neurostimulator, ON dopaminergic medication, with stimulation both ON or OFF.

Trait anxiety was measured with validated clinical scales in all participants, and state anxiety was measured with momentary assessment scales at multiple time points in the two at-home cohorts. Power in theta (4-8 Hz) and alpha (8-12 Hz) ranges were extracted from the LFP recordings, and their relation with anxiety ratings was assessed using linear mixed-effects models.

**Results:** In total, 33 PD patients (59 hemispheres) were included. Across three independent cohorts, with stimulation OFF, basal ganglia theta power was positively related to trait anxiety (all p<0.05). Also in a naturalistic setting, with individuals at home at rest with stimulation and medication ON, basal ganglia theta power was positively related to trait anxiety (p<0.05). This relationship held regardless of the hemisphere and DBS target. There was no correlation between trait anxiety and premotor cortical theta-alpha power. There was no within-patient association between basal ganglia theta-alpha power and state anxiety.

**Conclusion:** We showed that basal ganglia theta activity indexes trait anxiety in PD. Our data suggest that theta could be a possible physiomarker of neuropsychiatric symptoms and specifically of anxiety in PD, potentially suitable for guiding advanced DBS treatment tailored to the individual patient’s needs, including non-motor symptoms.

## Introduction

People with Parkinson’s disease (PD) experience a range of non-motor symptoms in addition to their cardinal motor symptoms, with non-motor symptoms having a significant impact on patients’ and caregivers’ quality of life ^1,2^. Among non-motor symptoms, neuropsychiatric deficits are prominent and can be present at any stage of the disease, even preceding motor deficits ^3^. Neuropsychiatric symptoms include, among others, depression, anxiety, apathy, psychosis, and impulse control disorders and related behaviors (ICB) ^4^. The prevalence of neuropsychiatric symptoms in PD is high, reaching up to 50% for depression and anxiety ^5^.

Anxiety has been recognized as one of the top three unmet therapeutic needs in PD patients and is recognized as a high research priority ^6^. Despite being so frequent, there are significant challenges in identifying, measuring, and treating anxiety in PD. People with PD, their caregivers, and physicians often lack awareness of the physical symptoms of anxiety and frequently attribute them to other medical conditions or to PD worsening ^7,8,9,10^. This leads to unnecessary investigations, hospital admissions, or increases in PD medications. The mechanisms underlying anxiety in PD are unclear and often multifactorial, encompassing multiple disease-specific and individual-specific factors, which complicates the development of effective treatments. Anxiety in PD can be persistent, episodic, or a combination of both, and anxiety can fluctuate during the day, influenced by dopaminergic medication intake ^11^. The temporal dimensions of anxiety are often captured in the constructs ‘trait’ anxiety - constituting an individual’s general level of anxiety over a considerable period, as opposed to ‘state’ anxiety, which reflects a person’s momentary and dynamic anxiety level ^12,13^. This etiological and temporal variability complicates the assessment of anxiety presence and severity, which currently relies on clinical interviews, clinician-administered scales, and self-reported questionnaires. This lack of objective measures of anxiety makes the management of this common symptom very challenging ^14,15^. Moreover, treatment options for anxiety in PD are currently limited, lack evidence-based support ^16^, and are often ineffective ^17^. Therapeutic strategies include pharmacological treatments (e.g. optimizing dopaminergic medication and antidepressants such as SSRIs and SNRIs) and non-pharmacological interventions (e.g. cognitive behavioral therapy) ^17^. Importantly, for patients with co-occurring anxiety and depression, effective treatment of depression does not necessarily alleviate anxiety, underscoring the dissociable nature of these symptoms as separable clinical entities in PD ^18^.

Altogether, anxiety management would greatly benefit from objective markers capable of indexing anxiety presence and severity, as well as principled and targeted treatments ^7^.

Deep brain stimulation (DBS) targeting either the subthalamic nucleus (STN) or globus pallidus pars interna (GPi) is an effective treatment for motor symptoms in PD ^19^ and may also improve non-motor symptoms ^20^, but the effect of DBS on neuropsychiatric symptoms and their fluctuations is still debated ^21^. Meta-analytic evidence suggests that, at the group level, anxiety symptoms improve after DBS ^22^. Electrophysiological activity such as local field potentials (LFPs) can be recorded from the STN or GPi during DBS surgery, immediately postoperatively through externalized electrodes, and via chronically implanted sensing-enabled neurostimulators ^23^. The latter even allows for exploring physiological markers in the naturalistic environment - the most relevant setting for PD patients. By providing direct access to subcortical activity, LFP research has offered valuable insights into disease mechanisms and cognitive functions, including the identification of physiomarkers of PD motor symptom states that enable precise adjustments of therapy via adaptive DBS ^24–26^. These increasingly available techniques provide a unique opportunity to identify objective physiomarkers of neuropsychiatric symptoms like anxiety that will be critical for understanding and treating non-motor symptoms in PD.

Whereas subcortical beta (13 – 30 Hz) power has been established as a physiomarker for PD motor symptoms ^27^, the neural correlates of neuropsychiatric manifestations of PD are less well-defined. Strikingly, although anxiety is such a common and disabling symptom, the neural correlates of anxiety using basal ganglia electrophysiology have never been assessed. For other neuropsychiatric symptoms, studies assessing the associations between these symptoms and subcortical neural signals have only been performed in the hospital setting, by recording intra- operatively or immediately after surgery from externalized DBS electrodes during periods that can be confounded by microlesion effects ^28,29^. Increased power in the theta-alpha (4-12 Hz) band has been suggested as a physiomarker of neuropsychiatric symptoms in PD ^30^. ICB ^31^ and depression ^32^ have been associated with increased STN theta-alpha and alpha power, respectively. The severity of trait impulsivity has been positively related to STN alpha power^33^. Other studies using behavioral paradigms have also implicated STN theta-alpha activity in emotional and behavioral processes ^30^.

In the present study, we aimed to investigate the relationship between intracranial theta-alpha activity and anxiety in three separate cohorts of PD patients with STN and GPi DBS, both in hospital and at home, using investigational and commercially available sensing-enabled neurostimulators.

## Material and methods

Three PD cohorts have been included and assessed in the present study (Fig. 1): (1) the ‘in-hospital externalized at-rest’ cohort: in-hospital LFP measurements via externalized DBS electrodes in patients with bilateral STN-DBS (London, UK), (2) the ‘at-home chronic at-rest’ cohort: at-home unsupervised LFP measurements through the chronically implanted Medtronic© Percept^TM^ PC neurostimulator in patients with STN-DBS and GPi-DBS (San Francisco, USA), and (3) the ‘at-home chronic task’ cohort: at-home supervised LFP measurements through the chronically implanted Medtronic© Summit RC+S neurostimulator in patients with STN-DBS and GPi-DBS performing a reward learning task (San Francisco, USA).

**Figure 1.**
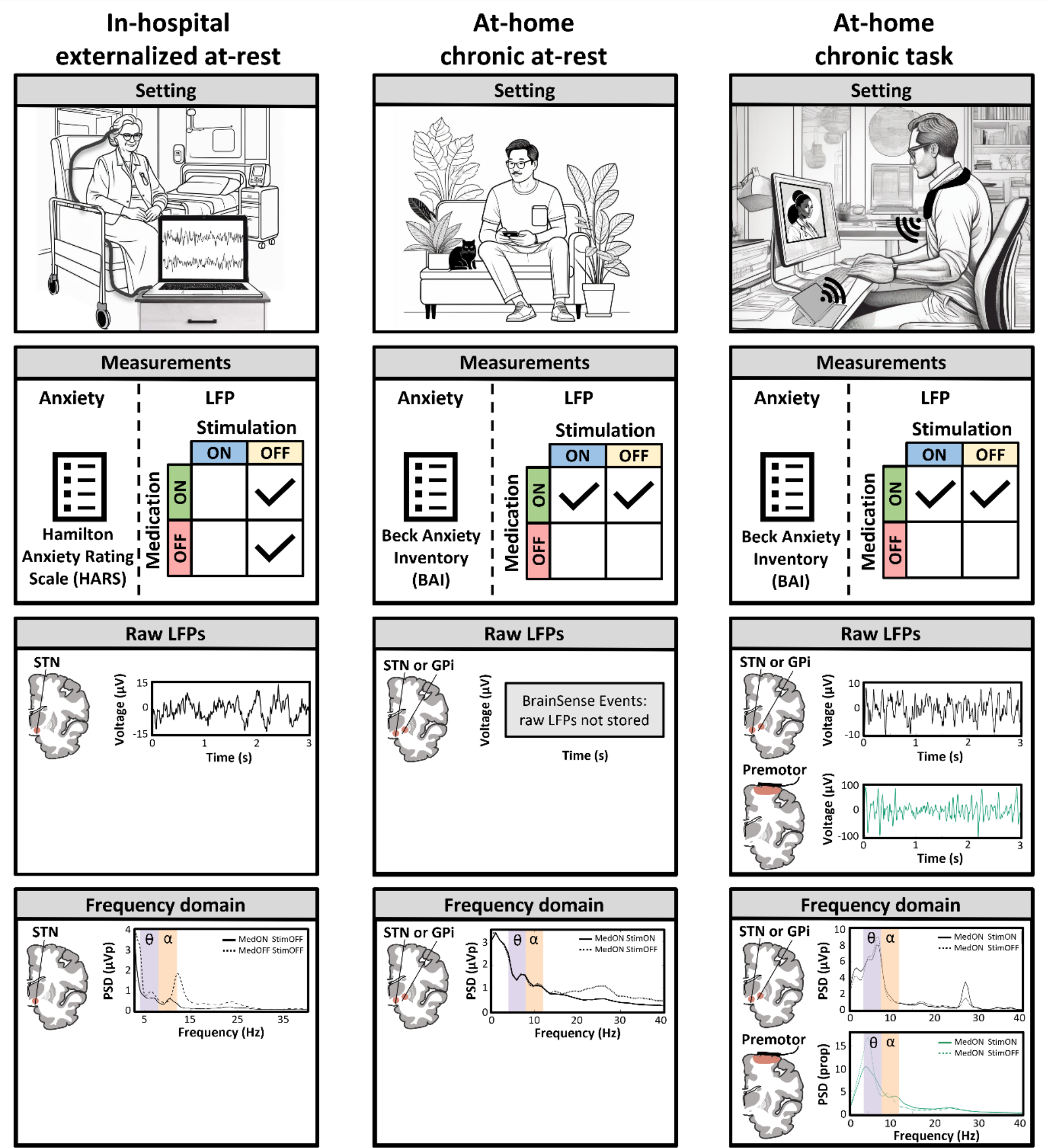
Three independent investigational cohorts for neurophysiological analysis. This study included three cohorts of PD patients treated with DBS. In the ‘in-hospital externalized at-rest’ cohort, basal ganglia LFP recordings were obtained from the STN a few days after surgery via externalized electrodes with the patient at rest. Patients were either OFF or ON medication, but stimulation was not enabled yet. Anxiety was rated with the Hamilton anxiety rating scale (HARS). In the ‘at-home chronic at-rest’ cohort, basal ganglia (STN or GPi) LFPs were obtained at home in an unsupervised but semi-structured setting using the Percept PC’s BrainSense technology (BrainSense ‘Event’ triggered by the patient) with the patient at rest. Stimulation was either OFF or ON, but patients were always in a Medication ON state. Anxiety was rated with the Beck anxiety inventory (BAI). Whereas in the other two cohorts offline LFP processing was required to obtain frequency domain data, raw LFPs were not available in the ‘at-home chronic at-rest’ cohort. In the ‘at-home chronic task’ cohort, basal ganglia (STN or GPi) and cortical (premotor cortex) LFPs were obtained via remotely supervised streaming via the Summit RC+S system with the patient at rest between trials of a behavioral task. Experimental conditions involved Medication ON Stimulation ON, and Medication ON Stimulation OFF. Anxiety was rated with the Beck anxiety inventory (BAI). In all three cohorts, analyses were focused on the relation between anxiety measures and power in the theta and alpha frequency bands. Infographics in the first row have been generated with the help of the generative AI platform Stable Diffusion Online (https://stablediffusionweb.com/). Abbreviations: LFP, local field potential; GPi, globus pallidus pars interna; PSD, power spectral density; STN, subthalamic nucleus.

### Patients

#### In-hospital externalized at-rest cohort

Consecutive PD patients (n = 12) undergoing bilateral STN-DBS were recruited at St. George’s University Hospital, London, United Kingdom (UK). Inclusion criteria were standard clinical criteria for DBS ^34^. In detail, they were: (1) diagnosis of idiopathic PD, (2) age < 70 years, (3) bothersome motor fluctuations and/or levodopa-induced dyskinesias despite optimal pharmacological management, (4) absence of dementia, major depression with suicidal thoughts or acute psychosis, (5) significant clinical response to levodopa challenge (at least 30% improvement in Movement Disorders Society-Unified PD Rating Scale III score), and (6) disease duration > 5 years.

#### At-home chronic at-rest cohort

Patients (n = 13) were recruited at the University of California San Francisco (UCSF), San Francisco, California, USA. Inclusion criteria were: (1) patients with idiopathic PD treated with unilateral or bilateral DBS of the STN or the GPi, (2) implanted with the Percept^TM^ PC neurostimulator, (3) on relatively stable DBS parameters and PD medication (i.e. beyond the DBS optimization phase - typically > 6 months after surgery), (4) stimulation parameters in at least one hemisphere compatible with BrainSense^TM^.

#### At-home chronic task cohort

Patients (n = 8) were recruited at University of California San Francisco (UCSF), San Francisco, California, USA. Patients were part of a clinical trial involving PD patients undergoing DBS implantation for motor fluctuations (NCT03582891). These patients were implanted with subcortical leads (Medtronic© models 3389 and 3387) in either the STN or the GPi, as well as quadripolar electrocorticography paddles (ECoG) (Medtronic© model 0913025) over the sensorimotor cortex. These electrodes were connected to an investigational sensing-enabled chronically implanted neurostimulator (Medtronic© Summit RC+S model B35300R ^35^).

### Data collection

#### In-hospital externalized at-rest cohort

Data were acquired 3 to 5 days after surgical insertion of the DBS leads, before the implantation of the neurostimulator while patients were admitted to hospital. Recording of STN LFPs (n = 24 hemispheres) was performed with a TMSi-Porti amplifier (TMS International, Oldenzaal, The Netherlands, fs = 2048 Hz) while participants were resting with eyes open, comfortably sitting on a chair for 5 minutes. Recordings were performed in two sessions on the same day; (1) in the morning after overnight withdrawal of antiparkinsonian medication (MedOFF condition) and (2) in the clinically defined ON medication condition, one hour after the administration of a participant’s regular dose of levodopa (MedON). The presence and severity of anxiety and depression were assessed using the Hamilton Anxiety Rating Scale (HARS) ^36^ and Hamilton Depression Rating Scale (HDRS) (Hamilton, 1960) ^36^, respectively on the day of the recording.

#### At-home chronic at-rest cohort

All participants were assessed in-hospital at baseline. During this visit, demographic and clinical data were gathered, and the presence and severity of anxiety and depression were rated using the Beck Anxiety Inventory (BAI) ^37^ and Beck Depression Inventory (BDI) ^38^ respectively. BrainSense^TM^ was enabled in the clinically active stimulation group on the Percept^TM^ PC neurostimulator. To this end, sensing was activated in a sandwiched configuration around the active stimulation contact, located in the STN (n = 14 hemispheres) or GPi (n = 6 hemispheres). Using the BrainSense^TM^ Event feature, an event called ‘Research’ was enabled for the patient. When an event is triggered by the patient using the patient programmer, a 30-second LFP recording is performed, the power-frequency spectrum of which is stored on the neurostimulator. Patients were instructed on the study protocol and how to use the patient programmer for the study at home. Patients performed research activities (see below) at home for a total of 14 days, in a structured but self-supervised manner. The timing of study activities was consistent within each patient and was in the medication ON state (i.e., 1 hour after taking a dose of levodopa). Patients performed two consecutive rounds of study activities on a research day: once with active stimulation at clinical amplitude (MedON StimON) and once with stimulation at 0.0 mA (MedON StimOFF). The order of these two activities differed across days, and was pseudorandomized across patients. One round of research activities involved the following consecutive activities: (1) setting the stimulation at clinical amplitudes or 0.0 mA using the patient programmer, (2) resting one minute, (3) triggering a ‘Research’ event using the patient programmer, (4) resting one minute, (5) rating current anxiety state on a visual analogue scale (VAS) where 0 is ‘not anxious at all’ and 100 is ‘very anxious’, (6) completion of a behavioral paradigm (to be reported separately). After completing the at-home research activities, data were downloaded (i.e. JSON file export) from the neurostimulator in the hospital at their next visit.

#### At-home chronic task cohort

Data were extracted from a previously conducted but unpublished study where participants performed a modified two-step reward learning task under remote supervision from the researcher while in their homes (see ^39^ for details of the task, which involves choices with fluctuating rewards to assess learning). Each participant performed this task between 9-13 times (n = 78 total sessions) across 5-7 days. VAS ratings of anxiety using the same scale as for the at-home at-rest cohort were collected immediately before starting the instruction phase of each task run. Experiments were performed in two clinical conditions: MedON StimON and MedON StimOFF, with at least 2 sessions per condition. Two patients could not tolerate the DBS therapy completely OFF, and were instead recorded at 53/50% and 92/92% of their clinical amplitude in the left/right hemispheres. LFP data were recorded during each task session subcortically from the pair of contacts surrounding the clinical stimulation contact (i.e., sandwich configuration) in the STN (n = 8 hemispheres) or GPi (n = 7 hemispheres) and cortically from bipolar pairs of the two most anterior and two most posterior ECoG contacts.

All ECoG contacts contributing to the cortical recording pairs reported here were located anterior to the central sulcus, ranging from the precentral gyrus to the middle/superior frontal gyrus. Posterior pairs were not analyzed due to variability in anatomical localization across subjects. At baseline, the presence and severity of anxiety and depression were evaluated using the BAI and BDI.

### Data analysis

#### In-hospital externalized at-rest cohort

All the acquired LFPs originated from a quadripolar electrode. Bipolar arrangement from the acquired data was obtained offline. In this way, each electrode presented three bipolar signals. Data were first inspected visually and parts of the signal with marked artefacts (e.g. signal saturation or movement artefacts) were removed (final signal length (mean ± standard deviation): 274.0 ± 77.5 s). Signals were then detrended and highpass filtered at 1 Hz (Butterworth). Spectral analysis was performed by computing the power spectrum of the signals using Welch’s method (2 s Hanning’s windows, 50% overlap) with a frequency resolution of 0.25 Hz. Theta and alpha bands were computed by averaging the band power between 4-8 Hz and 8-12 Hz, respectively.

#### At-home chronic at-rest cohort

The LFP data from the ‘Research’ events for the 14 days of at-home assessments were retrieved from the JSON files. Within each ‘Research’ event, LFP data were proportionally normalized per frequency band by dividing the sum of powers per canonical frequency band (i.e. theta 4- 8 Hz or alpha 8-12 Hz) over the sum of powers of the frequency range from 0 to 57.62 Hz. This latter upper limit was implemented because the powers above a varying frequency are censored by the processing onboard Percept PC, the cut-off of which depends on the stimulation frequencies (e.g. frequencies above 57.62 Hz are censored when stimulating at 180 Hz). For group-level comparisons of baseline BAI and BDI scores or average VAS anxiety with alpha or theta power, proportionally normalized powers per frequency band were averaged across events within hemispheres.

#### At-home chronic task cohort

LFP data during task sessions were reconstructed using the *processRCS* analysis toolbox ^40^. Data were extracted from 0.9 s epochs when participants were resting during the inter-trial interval. Trials were excluded for missing behavioral and/or neural data and practice trials (mean +/- SD: 840.6 +/- 165.0 trials/person). Power spectral density (PSD) estimates were extracted from these time series using Welch’s method (*pwelch* in MATLAB) with 0.4 s windows and 50% overlap for frequencies ranging from 1 to 55 Hz in 0.5 Hz steps. To eliminate differences between subjects and regions, the PSDs were then normalized for each trial by dividing by the sum of all power bands from 1-55 Hz. Normalized PSDs were then averaged across trials within each session before computing theta and alpha power as the mean within 4-8 Hz and 8-12 Hz bands, respectively. Theta and alpha power were then averaged across runs within each participant, yielding one estimate for each hemisphere within each participant.

### Statistical analyses

Because data were structured with a high level of non-independence, especially concerning patients with bilateral LFP recordings, linear mixed-effects (LME) models were used to assess the relation between the power of LFP frequency bands and behavioral measures (i.e. HDRS, HARS, BAI, BDI, and VAS anxiety). LME models predicted neural power averaged within each hemisphere of each patient using fixed effects of each participant’s symptom score, hemisphere (left/right), basal ganglia region (STN/GPi), and random intercepts for each participant. Separate models were used to predict either theta or alpha power using one of the a priori defined symptom metrics. One-sided likelihood ratio tests were used to assess the significance of the relationships by using the *compare* function in Matlab R2023a (Mathworks, Natick, MA), testing whether the model including the fixed effect of the symptom predictor of interest (e.g. BAI) improved model fit at a *p*-level defined at 0.05 compared to a null model without that predictor. The same LME model structure was used to assess state anxiety by using VAS anxiety ratings for individual events in the ‘at-home at-rest’ cohort and runs in the ‘at-home task’ cohort to predict low frequency power, again with separate models for theta and alpha power.

### Ethical approval

All patients provided written informed consent under the Declaration of the Principles of Helsinki. For the London cohort, approval was obtained from the Institutional Review Board of the Integrated Research Application System (IRAS) with the study protocol number 268941 - “Neural basis of neuropsychiatric symptoms in PD”. For the UCSF cohorts, approval was obtained from the Institutional Review Board of the University of California, San Francisco (UCSF) – study numbers 10-01350 and 20-31239 respectively.

## Results

In total, 33 participants (59 hemispheres) were included in this study; 12 participants in the ‘in-hospital externalized at-rest’ cohort, 13 in the ‘at-home chronic at-rest’ cohort, and 8 in the ‘at-home chronic task’ cohort. Demographics and clinical data per cohort are displayed in Table 1.

**Table 1.** Patient characteristics across the three cohorts. . Abbreviations: BAI, Beck anxiety inventory; BDI, Beck depression inventory; GPi, globus pallidus pars interna; HARS, Hamilton anxiety rating scale; HDRS, Hamilton depression rating scale; N/A, not applicable; SD, standard deviation; STN, subthalamic nucleus. *, each with a quadripolar electrocorticography paddle over the sensorimotor cortex

| Characteristics | In-hospital<br>externalized at-rest | At-home<br>chronic at-rest | At-home<br>chronic task |
| --- | --- | --- | --- |
| Patients (n) | 12 | 13 | 8 |
| Hemispheres (n) | 24 | 20 | 15* |
| Target (n hemispheres) - STN \| GPi | 24 \| 0 | 14 \| 6 | 8 \| 7 |
| Sex (n patients) - F \| M | 8 \| 4 | 2 \| 11 | 1 \| 7 |
| Age (y) - mean (SD) | 62.5 (3.9) | 64.9 (9.8) | 61.9 (9.6) |
| Disease duration (y) - mean (SD) | 11.2 (5.6) | 11.3 (4.2) | 11.4 (5.7) |
| DBS duration (y) - mean (SD) | N/A | 1.5 (1.8) | 2.5 (1.1) |
| Anxiety |  |  |  |
| *BAI - mean (SD)* | N/A | 9.9 (9.3) | 10.5 (3.5) |
| *HARS - mean (SD)* | 9.2 (7.6) | N/A | N/A |
| Depression |  |  |  |
| *BDI - mean (SD)* | N/A | 6.2 (3.7) | 9.8 (8.0) |
| *HDRS - mean (SD)* | 8.4 (5.4) | N/A | N/A |

First, we aimed to assess whether the low-frequency bands which have been implicated in neuropsychiatric/cognitive aspects of PD are also associated with anxiety using LFP recordings obtained shortly after surgery via externalized leads. In the ‘in-hospital externalized at-rest’ cohort, anxiety (as per HARS score) was positively related with STN theta power with MedOFF StimOFF (p = 0.031, β = 0.046), but not MedON StimOFF (p = 0.347, β = 0.061) (Fig. 2, panels A, B, and E). Anxiety was not related to STN alpha power in either condition (Fig. 2, panels C, D, and E).

**Figure 2.**
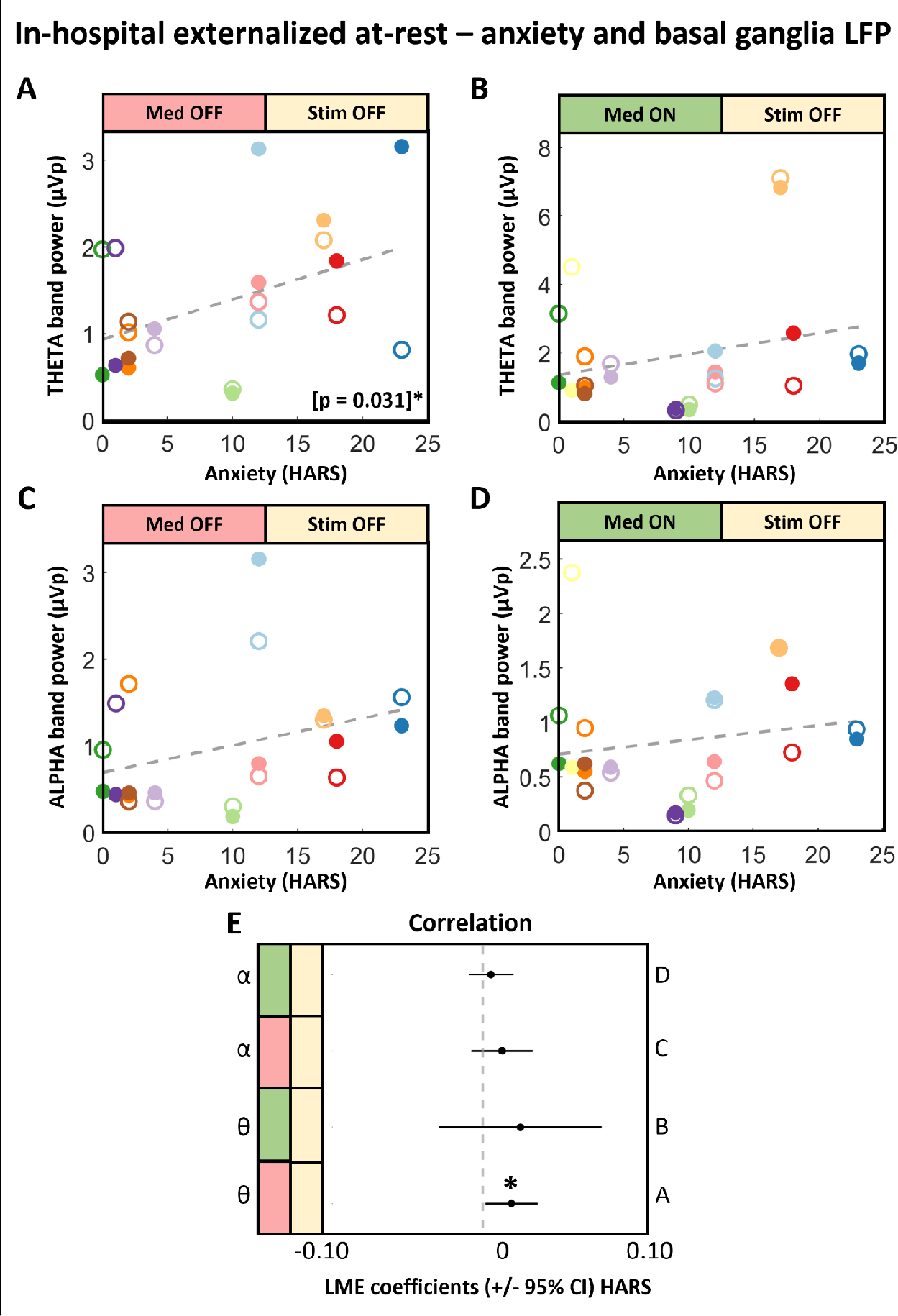
Anxiety is related to basal ganglia theta in in-hospital externalized recordings. This figure displays the relation between anxiety measured by the Hamilton anxiety rating scale and basal ganglia (STN) theta and alpha band power in the ‘in-hospital externalized at-rest’ cohort. Each color represents a patient (n = 12 patients), empty/filled markers indicate right/left hemisphere recordings (n = 24 hemispheres). With Medication OFF Stimulation OFF, a positive relation is present between anxiety and basal ganglia theta (panel A). Panel E illustrates the coefficients of anxiety in LME models. *, p < 0.05. Abbreviations: CI, confidence interval; HARS, Hamilton anxiety rating scale; LME, linear mixed-effects model; STN, subthalamic nucleus.

Next, we assessed whether the relation between anxiety and basal ganglia theta power holds during at-home LFP recordings of patients with chronically implanted sensing-enabled neurostimulators. In the ‘at-home chronic at-rest’ cohort - with unsupervised LFP recordings (average of 21.9 (SD 5.4) BrainSense^TM^ Event recordings per patients), anxiety (as per BAI score) was positively related to basal ganglia theta power in both the MedON StimOFF (p = 0.022, β = 0.0021) and MedON StimON (p = 0.022, β = 0.0016) conditions (Fig. 3, panels A, B, and E). Moreover, in this cohort, anxiety was also positively related to basal ganglia alpha power in both conditions (p = 0.012 and 0.020, β = 0.0021 and 0.0016) Fig. 3, panels C, D, and E). In the ‘at-home chronic task’ cohort - with supervised LFP recordings during a reward learning task, anxiety (as per BAI score) was positively related with basal ganglia theta power in the MedON StimOFF (p = 0.034, β = 0.0022) condition, and this relationship was trending towards significant with MedON StimON (p = 0.079, β = 0.0015) (Fig. 4, panels A, B, and E). Anxiety was not related to basal ganglia alpha power in any conditions for either at-home cohort (Fig. 4, panels C, D, and E).

**Figure 3.**
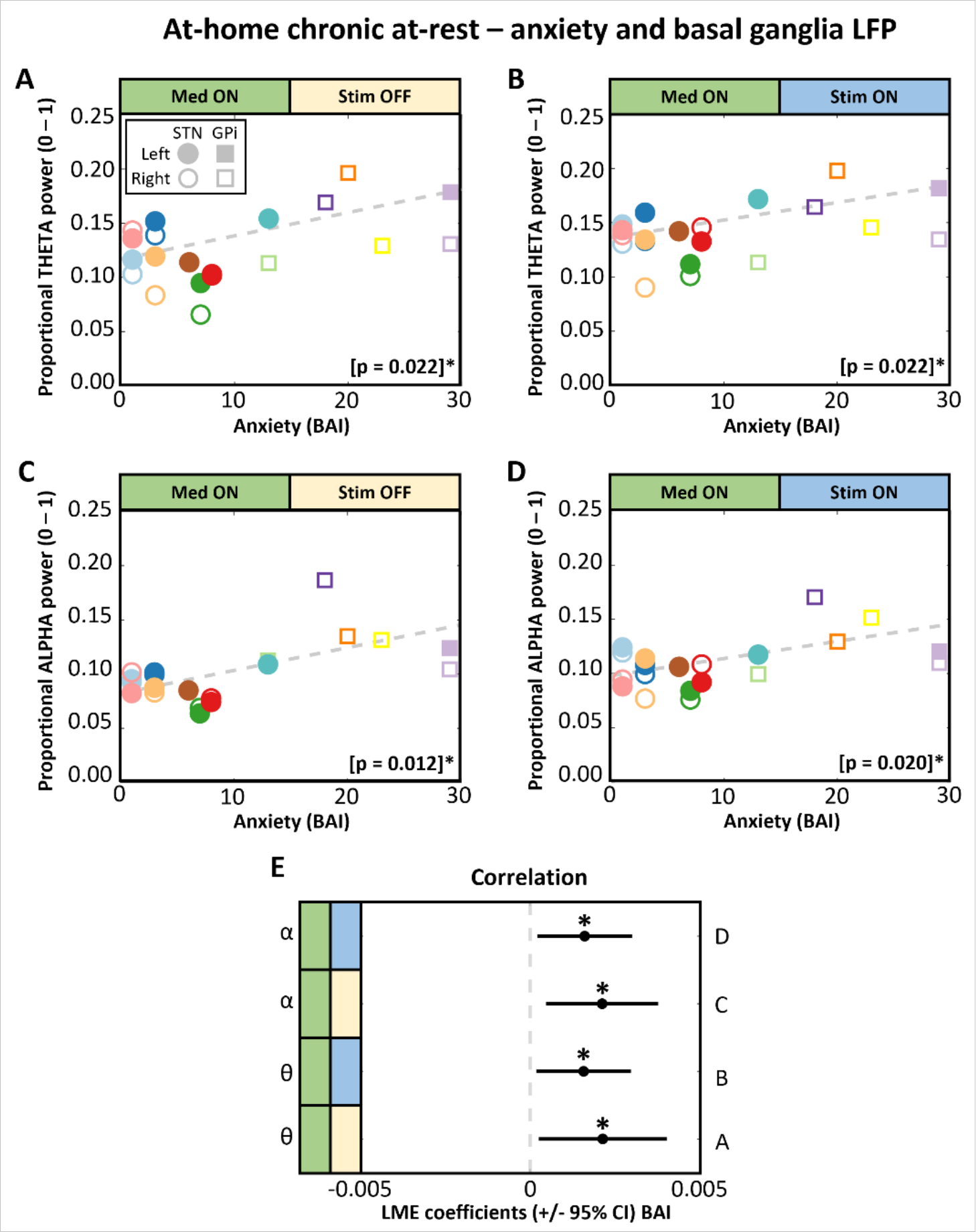
Anxiety is related to basal ganglia theta and alpha in at-home at-rest recordings. Relation between anxiety and subcortical LFP in the ‘at-home chronic at-rest’ cohort. Scatter plot colors indicate participant (n = 13 patients), with circles indicating STN and squares indicating GPi implants. Empty/filled markers indicate right/left hemisphere recordings (n = 19 hemispheres). Theta and alpha band power were extracted from at-home at-rest BrainSense Event LFP recordings, and correlation was assessed with baseline anxiety measured with Beck anxiety inventory (BAI). With Medication ON Stimulation OFF (panels A and C) and Medication ON Stimulation ON (panels B and D), anxiety is positively related with subcortical theta (panels A and B) and alpha (panels C and D) band power. Panel E illustrates the coefficients of BAI in LME models. *, p < 0.05. Abbreviations: BAI, Beck anxiety inventory; CI, confidence interval; GPi, globus pallidus pars interna; LFP, local field potential; LME, linear mixed-effects model; STN, subthalamic nucleus.

**Figure 4.**
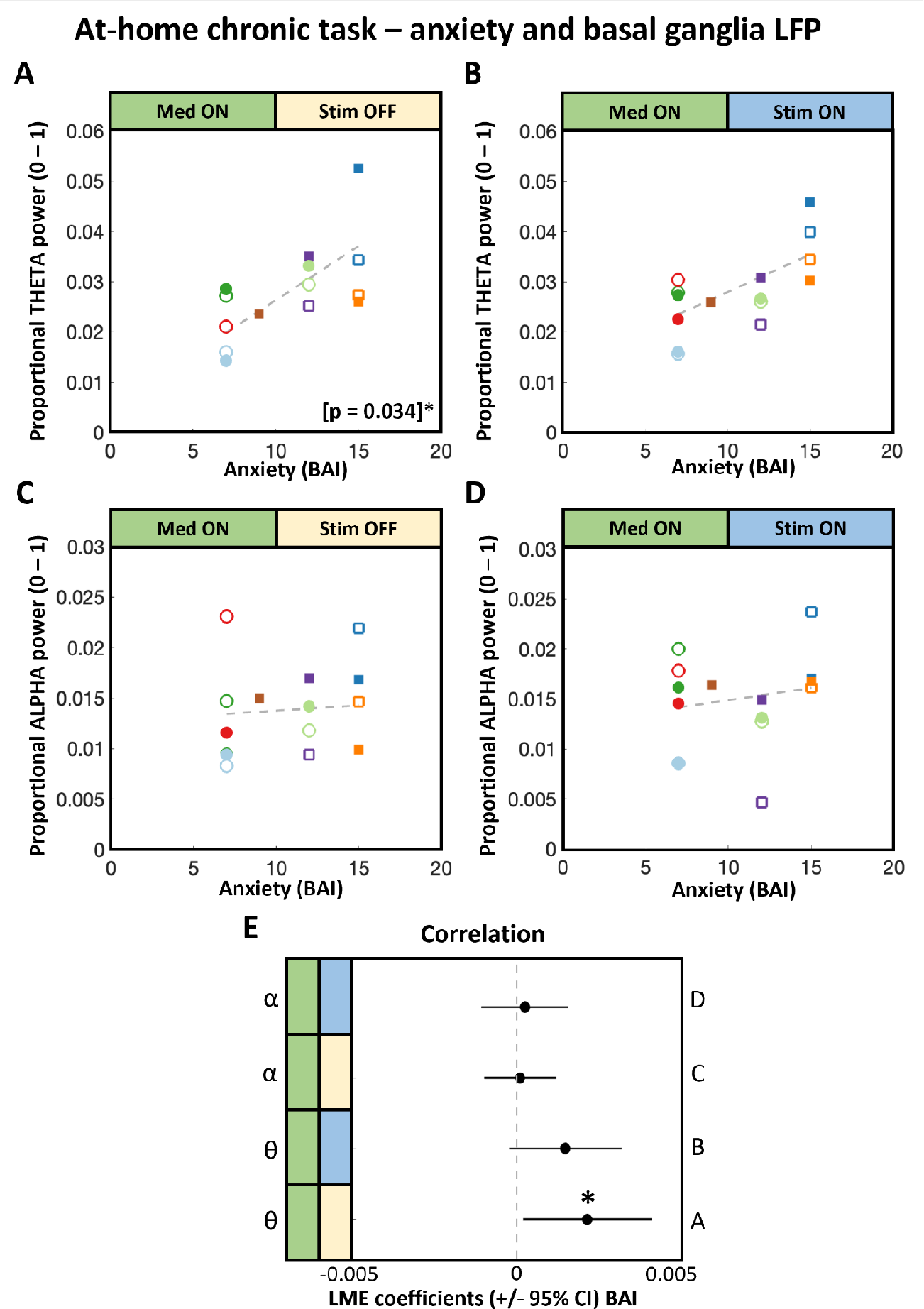
Anxiety is related to basal ganglia theta in at-home recordings during a cognitive task. Relation between anxiety and subcortical LFP during a reward task in the ‘At- home chronic task’ cohort. Theta and alpha power were extracted from the inter-trial interval in a reward task, and correlation was assessed with baseline anxiety (BAI). Scatter plot colors indicate participant (n = 8 patients), with circles indicating STN and squares indicating GPi implants. Empty/filled markers indicate right/left hemisphere recordings (n = 15 hemispheres). With Medication ON Stimulation OFF (panel A), there is a positive relation between anxiety and subcortical theta band power. Panel E illustrates the coefficients of BAI in LME models. *, p < 0.05. Abbreviations: BAI, Beck anxiety inventory; CI, confidence interval; GPi, globus pallidus pars interna; LFP, local field potential; LME, linear mixed-effects model; STN, subthalamic nucleus.

Altogether, a positive relation between anxiety and basal ganglia theta power was consistently present across three independent cohorts in both subacute clinical and naturalistic environments.

We further aimed to assess whether the relation between anxiety and theta is specific to the basal ganglia structures targeted with DBS (i.e. STN and GPi) compared to anatomically connected premotor cortical areas. In the ‘at-home chronic task’ cohort, electrocorticography data over the premotor cortex showed that anxiety was unrelated to cortical theta or alpha power (all *p* > 0.560, Supplementary Fig. 1).

Next, we aimed to investigate whether, beyond indexing ‘trait’ anxiety (e.g. HARS and BAI), basal ganglia theta power could also be related to within-subject variations in ‘state’ anxiety. This was assessed in the at-home cohorts. For the ‘at-home chronic rest’ cohort, for a period of approximately 14 days, patients provided daily reports of ecological momentary assessments of their anxiety level on a VAS immediately after triggering an LFP recording. At the within-subject level, VAS anxiety - representing ‘state’ anxiety - was not related to basal ganglia theta or alpha power, neither in MedON StimOFF (p = 0.147 and β < 0.0001 for theta, p = 0.832 and β < 0.0001 for alpha) nor MedON StimON (p = 0.960 and β < 0.0001 for theta, p = 0.870 and β < 0.0001 for alpha) conditions (Supplementary Fig. 2, panels A to E). Similarly, VAS anxiety ratings did not predict low frequency power in the ‘at-home chronic task’ cohort either. Specifically, VAS anxiety ratings were acquired immediately before starting the task to provide a measure of anxiety fluctuations across days, and as in the ‘at-home chronic at-rest cohort’, VAS anxiety scores at the run level did not predict either theta or alpha power in MedON StimOFF or MedON StimON (all p > 0.143, all β < 0.0001) conditions (Supplementary Fig. 2, panels F to J). This suggests that within-subject ‘state’ anxiety fluctuations are not directly related to basal ganglia theta power. In contrast, in the ‘at-home chronic rest’ cohort, averaging VAS anxiety scores per patient across 14 days to derive an estimate of ‘trait’ anxiety revealed a positive relationship with basal ganglia theta power in MedON StimOFF (p = 0.023, β = 0.0008) and was trending towards significant in MedON StimON (p = 0.051, β = 0.0007) (Supplementary Fig. 3, panels A, B, and E). Average VAS anxiety per patient was also positively related with basal ganglia alpha power, with MedON StimOFF (p = 0.012, β = 0.0009) and MedON StimON (p = 0.032, β = 0.0007)(Supplementary Fig. 3, panels C, D, and E). Thus, ‘trait’ measures of anxiety computed by averaging VAS anxiety ratings across days also predict theta and alpha power, which aligns with our core finding of a relationship between low frequency power and ‘trait’ anxiety measured by surveys (i.e. HARS and BAI). In sum, basal ganglia theta and alpha power reflected the average VAS anxiety ratings across days in the ‘at-home chronic at-rest’ cohort, but did not track day-to-day fluctuations in anxiety in either at-home cohort, indicating a stronger correspondence with ‘trait’ than ‘state’ anxiety levels.

Lastly, we explored the relationship between depression and low-frequency oscillations ^32^, but found inconsistent results across the three cohorts (See Supplementary Material).

In summary, our study revealed a consistent positive association between anxiety and basal ganglia theta power in PD patients across different cohorts and conditions, indicating a potential neural correlate of anxiety in PD.

## Discussion

In the present study, we evaluated the basal ganglia neural correlates of anxiety in PD patients for the first time, using both in-laboratory and at-home recordings. Across three independent cohorts, our results demonstrate that subcortical theta power recorded from STN and GPi is positively related to anxiety in PD, with higher theta power indexing more severe anxiety. This effect was present regardless of DBS being enabled/disabled and during both rest and waiting periods of a cognitive task. We did not find any association between anxiety and theta or alpha power recorded from the electrocorticography over the premotor frontal cortex, suggesting this relationship is region-specific. Lastly, theta did not track variations in anxiety ratings across days.

To the best of our knowledge, this is the first study exploring neural physiomarkers for non-motor symptoms using commercially available sensing-enabled DBS devices that allow chronic patient recording at home. Our approach of parallel lab-based and home-based assessments and recordings supports both scientific reproducibility and ecological validity.

Another feature of this study is the observation of the same relation between basal ganglia theta and anxiety in three independent cohorts, each employing somewhat different methodologies with regard to clinical setting, assessment of anxiety, and LFP recording. We studied a heterogeneous and inclusive DBS population comprising unilateral and bilateral DBS of both the STN and the GPi, with LME models accounting for heterogeneity across implantation targets and laterality.

Low frequency oscillations (including alpha and theta bands) in the STN have previously been evaluated in relation to cognition and emotion in PD patients (for a review see Ricciardi et al., 2023). Current literature indicates that low frequencies are implicated in cognitive processes such as conflictual judgement (Fumagalli et al., 2011), decision making (Rosa et al., 2013), reward-related processing (Zenon et al., 2016) and perceptual discrimination (Herz et al., 2016). In relation to neuropsychiatric symptoms, low frequencies have been related with trait impulsivity (Ricciardi et al., 2020), ICB (Alegre and Valencia, 2013; Rodriguez-Oroz et al., 2011), and depression (Brucke et al., 2007; Huebl et al., 2011; Kuhn et al., 2005).

We extend this knowledge to anxiety, showing a consistent association between theta and anxiety in each of our three samples. It is however not possible to disentangle whether the relationship with theta is strictly specific for anxiety or if it is driven by other highly correlated and co-occurring neuropsychiatric symptoms, especially depression ^41^. In our cohort of patients studied subacutely after surgery, theta was also positively related to depression, but importantly, this was not replicated in our two at-home cohorts. Overall, our data suggest that subcortical theta may specifically index anxiety and provide some evidence that anxiety and depression may be neurophysiologically separable. Nevertheless, this remains a challenge for further research since neuropsychiatric symptoms in PD are highly comorbid ^41^. A PD patient with anxiety is very likely to also present clinical symptoms of depression, ICB or apathy. Moreover, the comorbidity of anxiety and depression is highly influenced by a strong overlap in diagnostic criteria and assessment scales. This overlapping may be responsible for artifactually increasing comorbidity rates ^42^, and we cannot exclude that this influences our results and the previous reports. From a clinical perspective, although it has been well demonstrated that anxiety can occur independently from depression in PD ^41^, this conundrum may be less of a concern for current therapeutic strategies, as treatments for the different neuropsychiatric symptoms are often similar. For example, antidepressants and psychotherapy are validated treatments for anxiety, depression and apathy in PD ^17^. However, disentangling the different neuropsychiatric features in PD remains an important although challenging research question for future studies and a primary aim for developing new, effective, therapeutic approaches.

Recording LFPs via sensing-enabled DBS at home in the Percept and RC+S cohorts also enabled a comparison with daily VAS anxiety severity ratings, but we found no evidence that theta tracked these day-to-day variations in anxiety. The psychobiological substrates of fluctuations in anxiety captured by VAS ratings may differ substantially from standardized clinical questionnaires (e.g. BAI and HARS), which measure an average level of anxiety over the previous few weeks. The former is often referred to as ‘state’ anxiety, while the latter is conceptualized as ‘trait’ anxiety ^43,13^. It’s therefore conceivable that ‘trait’ and ‘state’ anxiety may have different physiomarker profiles. The lack of correspondence between theta and day-to-day anxiety VAS ratings, as well as the persistence of the relationship to clinical anxiety scores even during quiet attentive periods in the reward task, suggest that basal ganglia theta is more closely related to ‘trait’ anxiety.

The dissociation between state and trait anxiety in terms of physiomarkers might also reflect the heterogeneous phenomenology of anxiety in PD, which encompasses persistent and episodic anxiety or a combination of both. Also, factors such as non-motor fluctuations and situation-specific anxiety (e.g. fear of falling) are not really explored with the currently available questionnaires or ecological momentary assessment tools. This measurement gap reflects a need in future studies for more disease-specific measures to disentangle ‘state’ and ‘trait’ anxiety in PD, possibly including objective measures such as peripheral physiology (e.g. heart rate variability, skin conductance, etc.).

A limitation of this study is the relatively small sample size, which has limited our analysis. The sample sizes of the cohorts did not allow a full, data-driven exploration of potential physiomarkers. Instead, we relied on a priori testing of hypotheses concerning canonical theta and alpha frequency bands, which was informed by prior literature implicating these signals in cognitive and emotional processing ^30^. However, it is worth noting that despite smaller sample sizes per cohort, this was independently replicated across three different cohorts and significant within each cohort. Our LME modelling approach included fixed effects for hemisphere and basal ganglia region to control for these potential confounds, as well as random intercepts to account for the hierarchical nature of our repeated-measures data. However, these theoretically motivated decisions reduced statistical power to detect effects with our sample sizes, which may explain some inconsistencies across the three cohorts with regard to presence/absence of a relation in the different stimulation-medication conditions. For example, with medication ON stimulation OFF there is a significant relation between basal ganglia theta and anxiety in two out of three cohorts - with a trend (β = 0.061) in the same direction in the remaining cohort. Similarly, restricted statistical power due to sample sizes creates challenges for reliably disentangling the different contributions of hemisphere (left versus right) and target (STN versus GPi). Finally, we showed an association between theta and anxiety, however we cannot ascertain whether this reflects a primarily pathological or a secondary/compensatory process. For example, top-down communication in theta frequencies from medial prefrontal cortex to STN reflects cognitive control adjustments during uncertainty and punishment ^44–46^, and this medial frontal theta signal is stronger in individuals with higher trait anxiety ^47,48^. These data support the proposal that chronically increased anxiety may be related to excessive theta signaling of uncertainty and punishment ^49^, but the observational nature of our study precludes any inferences about brain-symptom causality and constrains our ability to assess theta’s role in the above mentioned dynamics of state anxiety.

Despite these limitations, the current study strongly suggests that basal ganglia theta could serve as an objective physiomarker of the general anxiety level in PD patients treated with DBS. This could have several clinical applications. First, subacute changes in basal ganglia theta (e.g. increased power over several weeks) may alert clinicians, caregivers and patients to be vigilant for anxiety symptoms. This could not only expedite and improve the precision of the diagnosis, but also could allow prompt initiation of anxiety-targeted therapies such as supportive measures, pharmacological and non-pharmacological interventions. Subsequently, basal ganglia theta may serve as a marker to assess and track response to these pharmacological and non-pharmacological interventions. Moreover, theta could inform DBS programming with regard to anxiety, similar to beta-based programming for motor symptoms ^50^. For example, theta physiomarkers could potentially identify DBS settings that may improve anxiety and could be used to monitor the effect of DBS on anxiety. Since DBS doesn’t have an acute effect on theta or generalized anxiety and our data suggest that theta indexes ‘trait’ anxiety (i.e. general level of anxiety), this theta-informed programming would likely operate on a timescale of weeks to months. Moreover, analogous to its effect on emotional processing and cognitive functions, delivering DBS at low stimulation frequencies (4 - 10 Hz, i.e., in the theta range) could be a strategy to improve the effect of DBS on anxiety ^51,52^. Finally, our work could potentially provide insights into the mechanism of similar neuropsychiatric symptoms in people who do not have PD. Indeed, alpha and theta frequencies power have been linked to major depression ^53^, drug addiction ^54^, obsessive-compulsive disorder ^55^ and Tourette syndrome ^56^, thus suggesting possible transdiagnostic networks.

More studies with a fully powered sample and rigorous methodology should confirm and extend our findings to clarify the causality of the relation between anxiety and theta in PD. More research is also required to assess whether, within a patient, theta changes along with long-term (weeks-months) changes in general anxiety level (‘trait’ anxiety). Analogous to beta-based adaptive DBS for motor symptoms ^24,25,26^, basal ganglia theta could drive closed-loop paradigms to provide principled responsive neuromodulation specifically targeted at anxiety. With our current findings, these would operate on longer timescales than beta-based motor aDBS. In order to develop a fast-responding closed-loop paradigm aimed to improve short-term fluctuations in anxiety, a physiomarker of ‘state’ anxiety will need to be established. Also, low frequency neural oscillations could be used to guide neurofeedback to train patients with PD to up and down-regulate their anxiety-related circuits and signals ^57^. Finally, the field would greatly benefit from the discovery of other objective markers of anxiety, residing in the subcortical/cortical LFP signal, autonomic system (e.g. heart rate variability ^58^, temperature, or respiration changes) or elsewhere.

In conclusion, we demonstrate that basal ganglia theta is related to anxiety in PD patients treated with DBS. This objective marker of anxiety could have diagnostic and therapeutic implications in clinical care, DBS programming, closed-loop DBS paradigms and neurofeedback protocols. Future research should further characterize theta and other potential objective physiomarkers of anxiety to eventually enable principled anxiety-targeted neuromodulation.

## Data availability

Individual patient data fall under the health data category of the General Data Privacy Regulation and require the lawful definition of data sharing agreements from all data controllers. Data sharing agreements can be set in place upon reasonable request to the senior authors (SL, LR).

## Acknowledgements

We thank our patients for participation in our studies. LR would like to thank Ioana Cociasu, Thomas Hart, Ilaria Bertaina and Rahul Shah for their help in collecting some of the data in the London cohort.

## Supplementary data

### Supplementary Results

#### Relationship between depression and low frequencies activity

In the ‘in-hospital externalized at-rest’ cohort, depression (as per HDRS score) was positively related to STN alpha and theta band power in the MedOFF StimOFF condition (p = 0.025 and β = 0.067 for alpha, p = 0.018 and β = 0.067 for theta), but not with MedON StimOFF (p = 0.716 and β = 0.034 for alpha, p = 0.439 and β = 0.019 for theta) (Supplementary Fig. 4, panel A). In the ‘at-home chronic at-rest’ cohort, depression (as per BDI) was positively related to basal ganglia alpha band power in MedON StimOFF (p = 0.031, β = 0.005), but not with MedON StimON (p = 0.620, β < 0.001) (Supplementary Fig. 4, panel B). In this cohort, depression was not related to basal ganglia theta band power (all p > 0.072, all β < 0.005) (Supplementary Fig. 4, panel B). In the ‘at-home chronic task’ cohort, depression (as per BDI) was not related to basal ganglia alpha nor theta band power in any of the conditions (all p > 0.05) (Supplementary Fig. 4, panel C).

## Supplementary Figures

**Supplementary figure 1.**
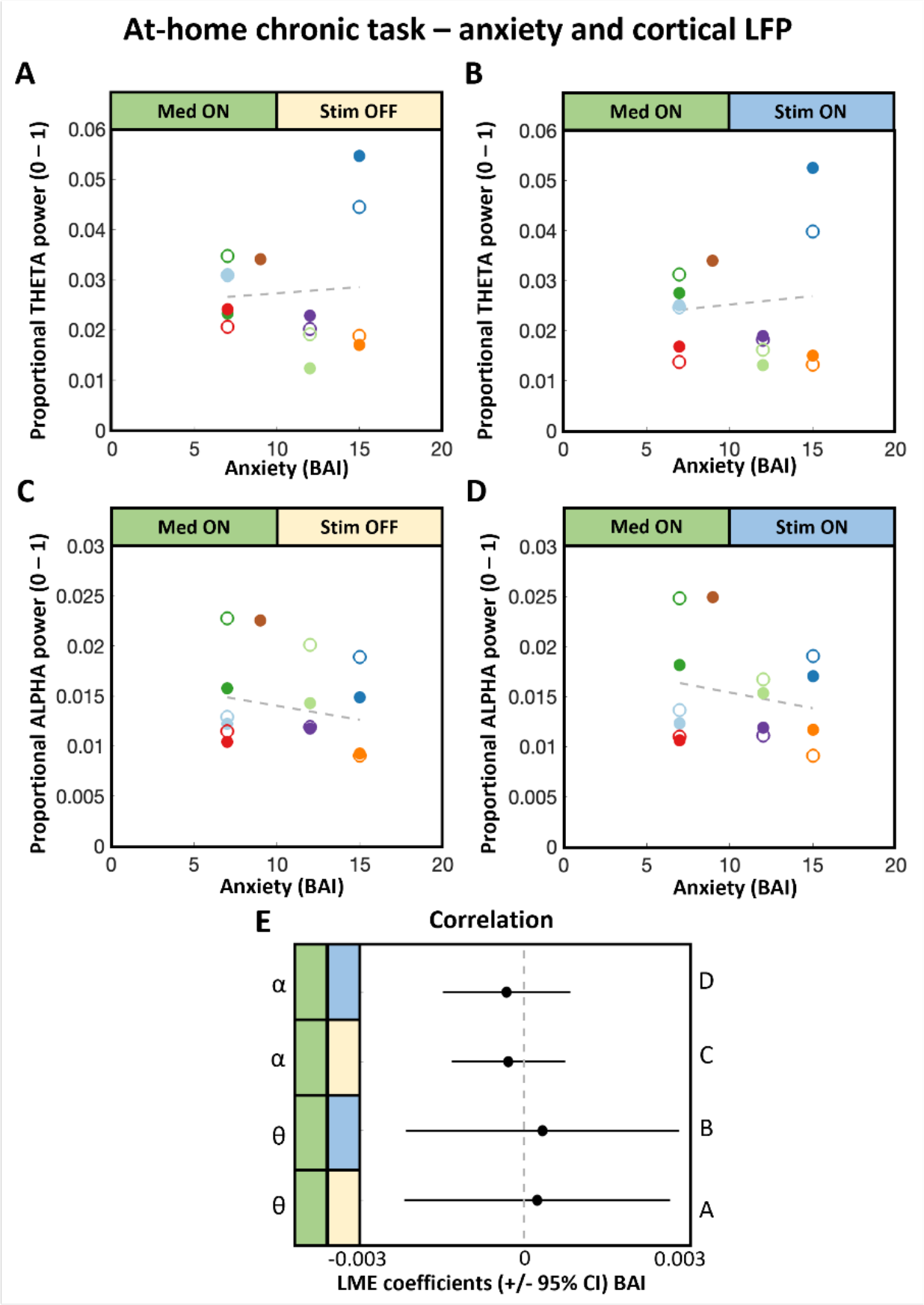
Anxiety is not related with premotor cortical theta or alpha. Relation between anxiety and premotor cortical LFP during a reward task in the ‘at-home chronic task’ cohort. Theta and alpha power were extracted from premotor electrocorticography recordings during the inter-trial interval in a reward task, and correlation was assessed with baseline anxiety (BAI). Scatter plot colors indicate participant (n = 8 patients), with circles indicating STN and squares indicating GPi implants. Empty/filled markers indicate right/left hemisphere recordings (n = 15 hemispheres). In none of the medication/stimulation conditions (*p* values for Panels A to D: 0.83, 0.77, 0.56, 0.56), anxiety is related to cortical theta or alpha band power. Panel E illustrates the coefficients of BAI in LME models. Abbreviations: BAI, Beck anxiety inventory; CI, confidence interval; LFP, local field potential; LME, linear mixed-effects model.

**Supplementary figure 2.**
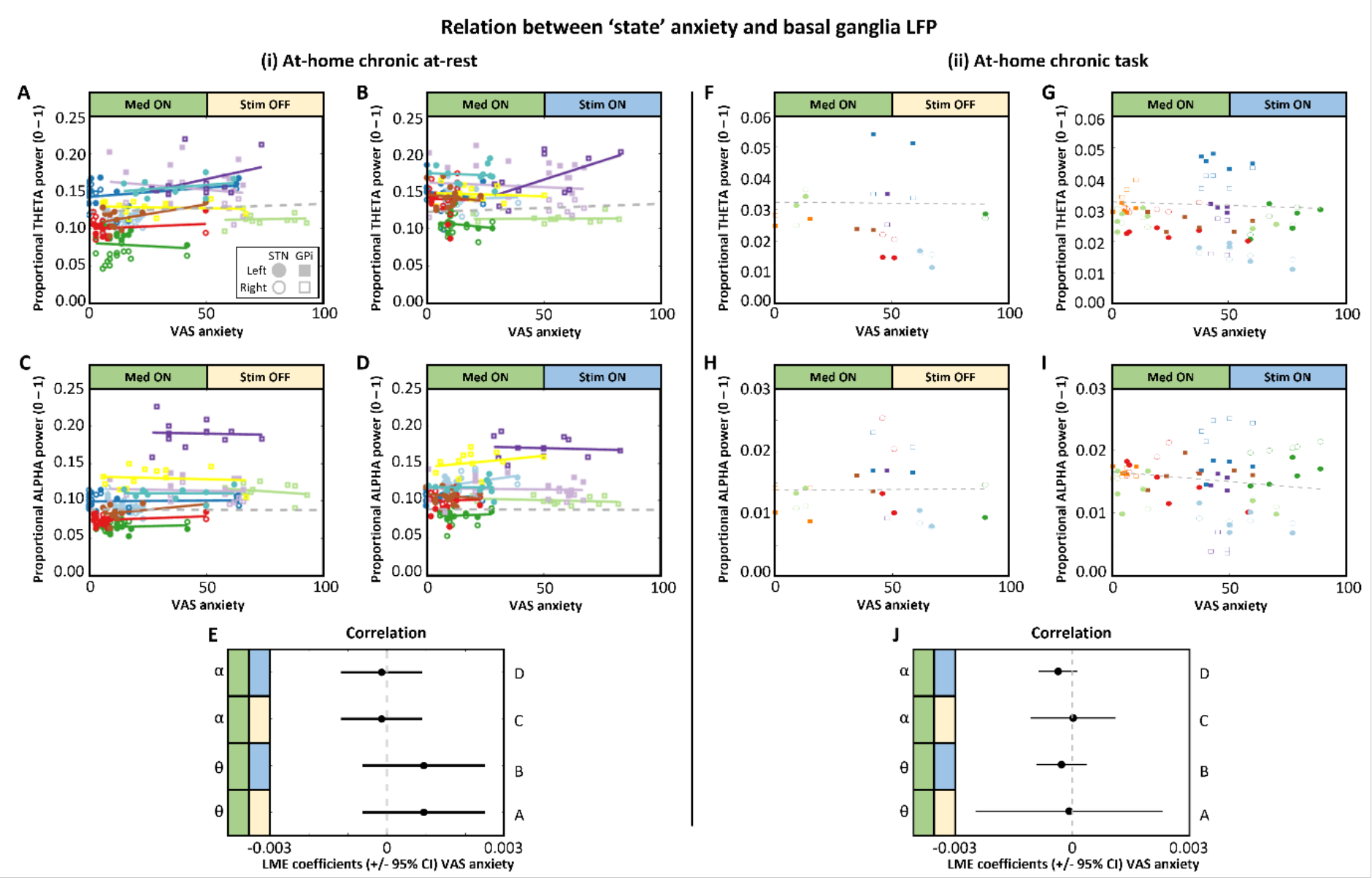
Basal ganglia theta and alpha do not track within-subject ‘state’ anxiety variation. Relation between state anxiety variation (as per VAS anxiety rating) and basal ganglia LFP in the ‘at-home chronic at-rest’ (panels A to E - n = 10 participants with at least five VAS ratings) and ‘at-home chronic task’ (panels F to J - n = 8 participants) cohorts. Scatter plot (panels A to D, and F to I) colors indicate participant, with circles indicating STN and squares indicating GPi implants, and empty/filled markers indicating right/left hemisphere recordings. Using an LME model, VAS anxiety was not related to basal ganglia theta or alpha in neither cohort (all p > 0.143, all β < 0.0001). Panels E and J illustrate the coefficients of VAS anxiety in LME models. Solid colored lines in panels A to D represent linear regression trendlines per participant - for visual representation purpose. Abbreviations: CI, confidence interval; LFP, local field potential; VAS, visual analogue scale.

**Supplementary figure 3.**
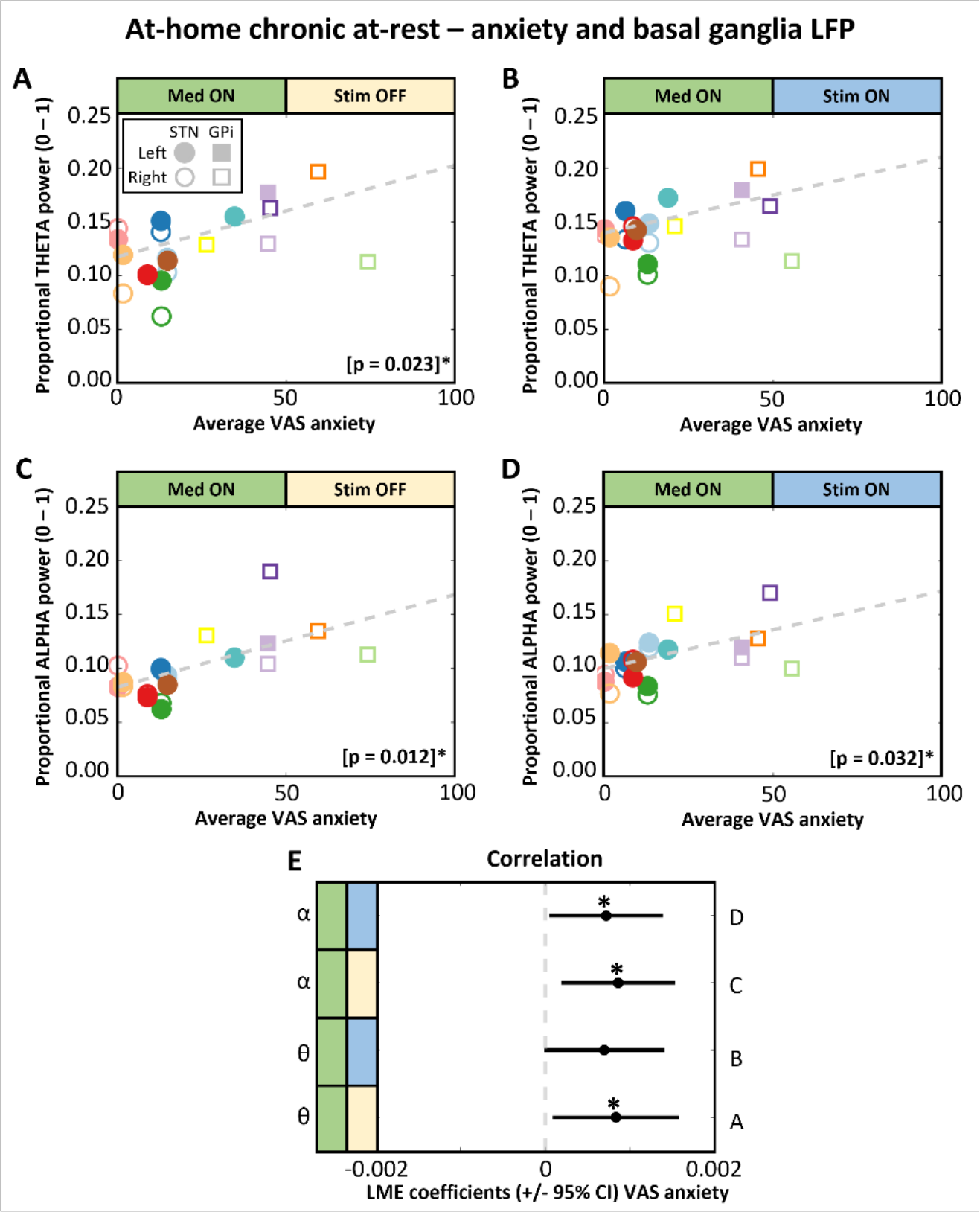
Average ‘state’ anxiety is related to basal ganglia theta and alpha in the at-home unsupervised environment. Relation between average state anxiety and subcortical LFP in the ‘at-home chronic at-rest’ cohort. Scatter plot colors indicate participant (n = 13 patients), with circles indicating STN and squares indicating GPi implants. Empty/filled markers indicate right/left hemisphere recordings (n = 20 hemispheres). Per hemisphere, theta and alpha band power were extracted from at-home at-rest BrainSense Event LFP recordings. Correlation was assessed with the, per patient, average momentaneous anxiety level measured over 14 days via a visual analogue scale (VAS). With Medication ON Stimulation OFF, anxiety is positively related with subcortical theta (panel A) and alpha (panel C) band power. With Medication ON Stimulation ON, anxiety is positively related with subcortical alpha (panel D) band power. Panel E illustrates the coefficients of BAI in LME models. *, p < 0.05. Abbreviations: CI, confidence interval; GPi, globus pallidus pars interna; LFP, local field potential; LME, linear mixed-effects model; STN, subthalamic nucleus; VAS, visual analogue scale.

**Supplementary figure 4.**
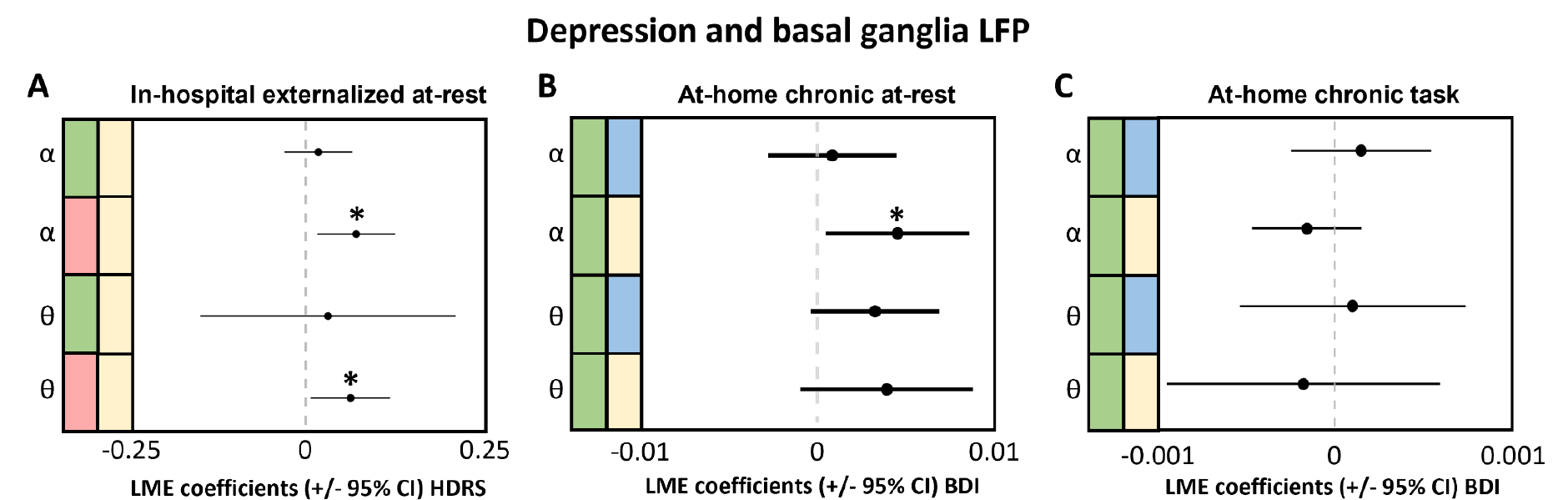
Relation between depression and basal ganglia LFP across three cohorts. Each panel illustrates the coefficients of depression (as per HDRS or BDI) in LME models assessing the relation between depression and basal ganglia LFP within each cohort. With medication and stimulation OFF, basal ganglia theta and alpha band power are positively related with depression (as per HDRS) in the ‘in-hospital externalized at-rest’ cohort (panel A). With medication ON and stimulation OFF, basal ganglia alpha band power is positively related with depression (as per BDI) in the ‘at-home chronic at-rest’ cohort (panel B). In the ‘at-home chronic task’ cohort, depression is not related to basal ganglia theta or alpha power with medication ON and stimulation ON or OFF (panel C). *, p < 0.05. Color legend: green, Medication ON; red, Medication OFF; blue, Stimulation ON; yellow, Stimulation OFF. Abbreviations: BDI, Beck depression inventory; CI, confidence interval; HDRS, Hamilton depression rating scale; LFP, local field potential; LME, linear mixed-effects model.

